# Digital risk score sensitively identifies presence of α-synuclein aggregation or dopaminergic deficit

**DOI:** 10.1101/2024.09.05.24313156

**Authors:** Ann-Kathrin Schalkamp, Kathryn J Peall, Neil A Harrison, Valentina Escott-Price, Payam Barnaghi, Cynthia Sandor

**Affiliations:** Division of Psychological Medicine and Clinical Neuroscience, School of Medicine, Cardiff University; Cardiff, United Kingdom; UK Dementia Research Institute, Cardiff University; Cardiff, United Kingdom; Neuroscience and Mental Health Innovation Institute, Division of Psychological Medicine and Clinical Neurosciences; Cardiff, United Kingdom; Cardiff University Brain Research Imaging Centre (CUBRIC); Cardiff, United Kingdom; UK Dementia Research Institute, Care, Research and Technology; London, United Kingdom; Department of Brain Sciences, Faculty of Medicine, Imperial College London; London, United Kingdom

## Abstract

**Background:** Use of digital sensors to passively collect long-term offers a step change in our ability to screen for early signs of disease in the general population. Smartwatch data has been shown to identify Parkinson’s disease (PD) several years before the clinical diagnosis, however, has not been evaluated in comparison to biological and pathological markers such as dopaminergic imaging (DaTscan) or cerebrospinal fluid (CSF) alpha-synuclein seed amplification assay (SAA) in an at-risk cohort.

**Methods:** To address this, we performed a cohort study using longitudinal clinical assessment data from the Parkinson’s Progression Marker Initiative (PPMI) cohort collected between 2010 and 2020 with additional long-term (mean: 485 days) at-home digital monitoring data (collected 2018-2020) from the Verily Study Watch. We derived a digital risk score and evaluated it in an at-risk cohort (N = 109) consisting of people with genetic markers (*LRRK2, GBA*) or prodromal symptoms (hyposmia, polysomnography-proven Rapid-Eye-Movement behavioral sleep disorder) without a diagnosis of PD for whom all modalities were available (digital, DaTscan, SAA). The digital risk score was compared to the Movement Disorder Society (MDS) research criteria for prodromal PD, alpha-synuclein SAA and DaTscan.

**Findings:** In the at-risk cohort (N=109, mean age = 64.62±6.86, 37% male), the digital risk correlated with the MDS research criteria for prodromal PD (r = 0.36, p-value = 1.46x10^-4^) and was increased in individuals with subthreshold Parkinsonism (UPDRS III > 6) (p-value = 4.99x10^-6^) and hyposmia (p-value = 3.77x10^-2^). Notably, the digital risk was correlated with DaTscan putamen binding ratio (r = -0.32, p-value = 6.64x10^-4^) and CSF SAA (r = 0.2, p-value = 3.9x10^-2^). The digital risk achieved higher sensitivity in identifying people with SAA positivity (0.71 vs 0.43) or DaTscan positivity (0.43 vs 0.14) than the MDS prodromal score but performed on-par or worse than hyposmia (SAA+: 0.71 vs 0.71, DaT+: 0.48 vs 0.57).

**Interpretation:** A digital risk score from smartwatch data could be used as a sensitive screening tool for early detection of PD followed by more specific tests.

## INTRODUCTION

The diagnosis of Parkinson’s disease continues to rely on clinical judgement, requiring evidence of motor signs. However, by this point 50-70% of the dopaminergic neurons will have degenerated (1). Therefore, identifying people prior to this is of high clinical value, and essential for the investigation of neuroprotective therapies.

Existing risk scores trying to identify individuals during the prodromal phase, such as the MDS research criteria (2) or PREDICT-PD (3), are based on lifestyle and genetic factors as well as prodromal symptoms. Such prodromal risk scores, however, show low sensitivity over ten-year follow-up (35%) (4, 5). Biological and pathological markers for PD have shown promising performance in prodromal cohorts, with DaT binding found to be reduced in ∼40% of patients with idiopathic RBD (6, 7) with 36.48% converting to PD within 4.7 years (8). Recently, CSF alpha-synuclein SAA has been found to be highly predictive for idiopathic PD, and to be present in individuals with hyposmia (88.9%) and RBD (84.8%) (9). Based on these markers, two biological definitions of PD have been suggested (10, 11). Despite the high specificity of these tests, they are not suited for population-based screening due to their associated cost, invasiveness, and time requirements.

Previously, we have shown that one week of accelerometer data can identify people years prior to their clinical diagnosis (12). Digital sensor data can be passively collected at home with low-cost devices, addressing the limitations of the above-mentioned tests. The relationship between biological measures of PD and digital risk has not yet been investigated, with positive findings underlining the validity of digital screening.

Here, we use the at-risk cohort without known phenoconversion from the deeply-phenotyped Parkinson’s disease Progression Marker Initiative (PPMI) (13) to compare a long-term digital risk score to established prodromal, pathological, and biological markers. For this, we 1) used a 1.3-years observation period, 2) incorporated data from a multi-sensor smartwatch, and 3) associated this digital risk not only with prodromal symptoms such as RBD and hyposmia, but also DaTscan and CSF alpha-synuclein SAA.

## MATERIALS AND METHODS

### Study Cohort

PPMI has collected data from those with recently diagnosed PD, people at risk, and unaffected controls since 2010. All 48 participating PPMI sites all received approval from an ethical standards committee before study initiation and written informed consent was obtained for all individuals participating in the study. The study was registered at clinicaltrials.gov (NCT01141023). This analysis used DaTscan and alpha-synuclein SAA results for prodromal participants, obtained from PPMI upon request after approval by the PPMI Data Access Committee. Between 2018 and 2020 a subset of participants has been supplied with a Verily Study Watch, which is equipped with a multitude of sensors including accelerometer, gyroscope, electrocardiogram, and photoplethysmography. We used the analytic dataset cohort assignment. The prodromal group was formed of people with polysomnography (PSG)-proven RBD, hyposmia, or mutations in Mendelian inherited genes considered causative or contributing to increased risk in PD (e.g. *LRRK2, GBA, SNCA, Parkin, Pink1*).

### Digital Data

Derived measures were provided by Verily (STable 1) and accessed in November 2022. The derived data including 1-hour interval timeseries data on sleep (total time, REM time, NREM time, deep NREM time, light NREM time, wake after sleep onset (WASO), awakenings, sleep efficiency), physical activity (step count, walking minutes), and vital signs (pulse rate, mean root mean squared successive differences (RMSSD) (heart beat), median RMSSD, RMSSD variance) was available for 149 PD cases, 158 prodromal cases, and 35 healthy controls covering a mean of 485 days. Six participants originally assigned to the prodromal group received a diagnosis of PD after recruitment, but before digital data collection, these were excluded from analysis.

### Clinical and biological data

Data was downloaded from PPMI in 2021 and access to sequestered data was provided in 2023. The following clinical assessments were retrieved: University of Pennsylvania Smell Identification Test (UPSIT) (14), UPDRS scores (15), Scales for Outcomes in Parkinson’s disease (SCOPA) autonome, and REM sleep behavior disorder screening questionnaire (RBDSQ) (16).

The most recent minimum putamen SBR data was calculated from DaTscan and were on average 0.33±1.91 years before the final available date of digital data collection.

CSF alpha-synuclein SAA data from baseline CSF samples included the mean Fmax values across the three repetitions and the SAA classification. These measurements were on average 3.4±1.38 years before the final date of available digital data. DaTscan and alpha-synuclein SAA results for prodromal participants were obtained after approval by the PPMI Data Access Committee.

### Prodromal markers and risk factors

We retrieved all data necessary to calculate the prodromal risk score from the MDS research criteria(2) (STable 2). We were not able to retrieve information on substantia nigra hyperechogeneicity or urate levels.

### Statistical Analysis

All analyses were performed in python v3.9 using sklearn 1.2.1 (17) for model training and evaluation, tsfresh 0.20.0 (18) for timeseries feature extraction, scipy 1.10.0 and pingouin 0.5.3 for statistical testing (19), and matplotlib 3.6.3 and seaborn 0.12.2 for creating figures. Data loading and manipulation has been facilitated through an adapted version of pypmi (https://github.com/rmarkello/pypmi). All associated code will be made available at https://github.com/aschalkamp/DigitalPPMI, which can be used to replicate the performed analyses and retrieve the digital risk score. Analysis and reporting followed the TRIPOD+AI guidelines.

### Digital timeseries feature extraction

First, the overall mean over time was computed for each subject for each digital marker. The PD group was significantly younger than the at-risk group (Cohen’s d = 0.51, p-value = 1.4x10^-5^) (STable 3) thus linear models were fit on the healthy controls (N = 35) to identify the effect of age and sex on each marker. The residuals were compared with two-sided T-tests with significant results defined as passing 0.05 FDR correction.

Second, tsfresh was applied for each individual for each raw digital feature to extract timeseries features. This resulted in 783 features such as maximum, minimum, skewness, kurtosis, and trend for each of the 14 digital timeseries, leading to a total of 10962 features per subject (STable 4).

### Risk models

The model described in the MDS research criteria(2) was implemented. We include two versions of this: 1) a full version, 2) a restricted version excluding DaTscan positivity. The latter serves as a better representation when applied in the general population.

We computed the digital risk score by training elasticnet logistic regression models identifying PD (N=143) from healthy controls (N=34) based on the computed digital timeseries features (STable 4). The at-risk group was not used for model training or validation at any point. A nested cross-validation was used with an inner and outer five-fold stratified split. Based on the training set, the data was standardised. The inner split was used to run a grid search to identify the best penalty parameter between 10^1^ and 10^4^ for alpha and between 0 and 1 for the L1 ratio (STable 5). The area under the precision-recall curve (AUPRC) was used as the evaluation score and the final model comparison. We compared its performance to a baseline model only using age, male sex, and education years as predictors. The predictors were assessed for stability and significance across folds considering multiple comparisons. The predicted probabilities were retrieved for all subjects, including the external test set of at-risk subjects, as the average over the outer folds.

The optimal threshold for identifying PD from healthy controls in terms of F1-score was found to be 0.54. We performed additional analyses on the effect of the considered timeframe, the considered feature sets, and the applied machine learning model on the performance (SMethods 1).

### Comparison of predicted risks

109 subjects in the at-risk group who did not yet phenoconvert and where future phenoconversion status is unknown had complete data available (STable 6). We compared the risk scores, pathological, and biological markers by correlating each pair using Pearson’s correlation. Significant correlations are reported when passing 0.05 FDR correction. We assessed which known prodromal markers and risk factors were associated with higher digital risk scores using Welch’s two-sided T-tests with 0.05 FDR correction. We only included markers for which 10 or more cases and controls were available (N=14). We further compared the estimated risk scores between the groups defined by the biological NSD staging system (10) and the biological SynNeurGe classification system (11).

### Evaluation of risk scores

Assuming that DaTscan or CSF SAA serve as the gold-standard for future conversion to PD, we assessed the performance of the digital risk model, the MDS prodromal model(2), and hyposmia in this scenario, computing recall, precision, and F1 score. We further assessed how a chaining of tests (i.e. first performing a digital screening and then sending all predicted positives to further tests) affected these performance metrics.

### Assessment of individuals with biological or pathological markers and low digital risk

We investigated which features were altered in those individuals not identified by the digital risk model but that had positive CSF SAA (N=4) or positive DaTscan (N=4). We computed Welch’s two-sided t-test for the maximum ever recorded UPDRS III score comparing individuals not identified by the digital risk with individuals showing both high biological/pathological risk and high digital risk. We repeated this analysis for hyposmia and the restricted MDS prodromal risk score and report results as significant when passing 0.05 Bonferroni correction.

## RESULTS

### Digital outcome measures capture changes in at-risk groups

The boxplots show the residual overall mean of digitally tracked sleep efficiency adjusted for age and sex with parameters learned from a linear regression on the healthy controls. The overall mean is computed over the whole observation time per subject for each group. The boxplots depict the group median and quartiles per group with the whiskers showing the Q3+1.5 interquartile range (IQR) and Q1-1.5 IQR (Parkinson’s disease cases: PD; healthy controls: HC; carriers of genetic risk alleles or prodromal symptoms without a diagnosis of PD: GBA, LRRK2, hyposmia, polysomnography-proven RBD, positive DaTscan, positive SAA; union of these: at-risk). The number in the yellow box indicates the number of individuals per group. Group differences were calculated with two-sided T-test comparing PD and HC to each of the at-risk groups. Lines and numbers show significant differences with 0.05 FDR corrected p-values.

We computed the mean for each digital measure over the complete period of observation. Comparing these average measures demonstrated that all physical activity measures were reduced in PD compared to controls (Figure 1, STable 7): step count (Cohen’s d = 1.24, p-value = 9.57x10^-10^), walking minutes (Cohen’s d = 1.04, p-value = 2.03x10^-7^). Four of the eight sleep measures were also significantly lower in individuals diagnosed with PD compared to controls; sleep length (Cohen’s d = 0.62, p-value = 2.56x10^-3^), sleep efficiency (Cohen’s d = 1.15, p-value = 1.69x10^-8^), REM (Cohen’s d = 1.2, p-value = 4.11x10^-9^), and deep NREM sleep length (Cohen’s d = 0.94, p-value = 3.37x10^-6^). None of the four digital vital signs were significantly different between PD cases and healthy controls.

**Figure 1:**
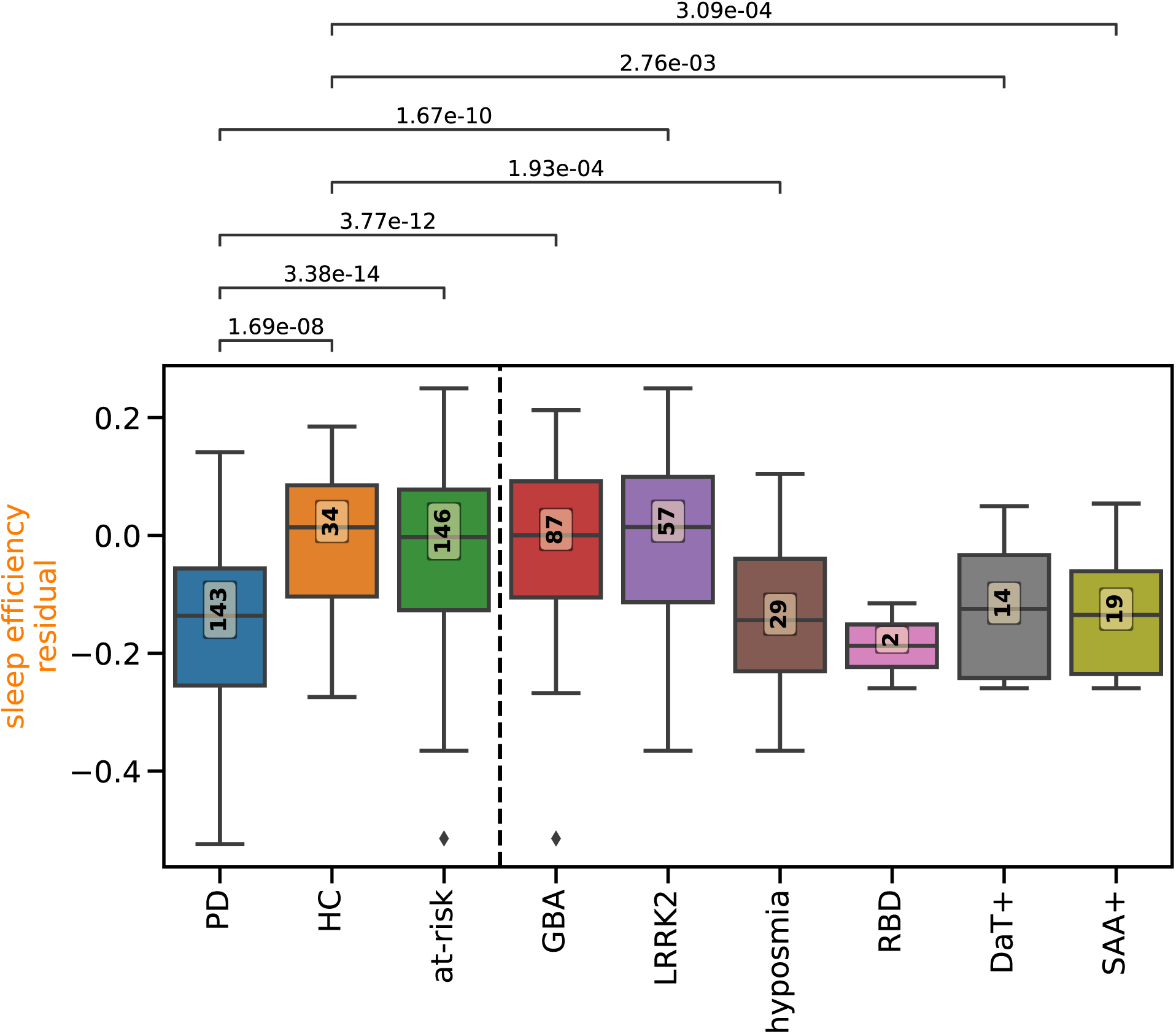
Digital measures capture differences in at-risk groups

Due to the heterogeneity of the at-risk group, we split the group by specific identifiers. The whole at-risk group including genetic carriers and those with prodromal symptoms, did not show any differences to healthy controls. Differences between controls and the at-risk subgroups were observed for sleep efficiency, which was significantly reduced in individuals with hyposmia (Cohen’s d = 1.04, p-value = 1.93x10^-4^), positive DaTscan (Cohen’s d = 1.05, p-value = 2.76x10^-3^), and positive SAA (Cohen’s d = 1.14, p-value = 3.09x10^-4^) (Figure 1). The hyposmia group showed significant differences to the controls in five measures, with SAA positivity showing differences in four, and DaTscan positivity in one measure (Figure 1, STable 7). These results highlight the heterogeneity of the at-risk cohort in PPMI where carriers of genetic mutations showed no difference to healthy controls and those with prodromal symptoms did differ in both physical activity measures, three sleep measures, and one vital signs measure (Figure 1, STable 7). Overall, the digital measures hold information relevant to PD and some of the associated prodromal markers.

### Long-term digital risk score relates to MDS prodromal risk and putamen binding ratio

We obtained digital risk scores (Figure 2) from models trained on the timeseries features for each of the 14 digital measures. The logistic regression model was trained to identify PD (N = 135) from healthy controls (N = 34), leaving the at-risk group (N = 109) as an external dataset not seen during training. The digital risk model (AUPRC = 0.96±0.01) significantly outperformed the baseline model (AUPRC: 0.8±0.04, p-value = 8x10^-6^). Consistently selected features predominantly originated from REM sleep time (41.39%) and step count (48.28%) (Supplemental Figure 2). Additional analyses on machine learning model, considered feature set, and included timeframe can be found in Supplemental Material (Figures 3-5).

**Figure 2:**
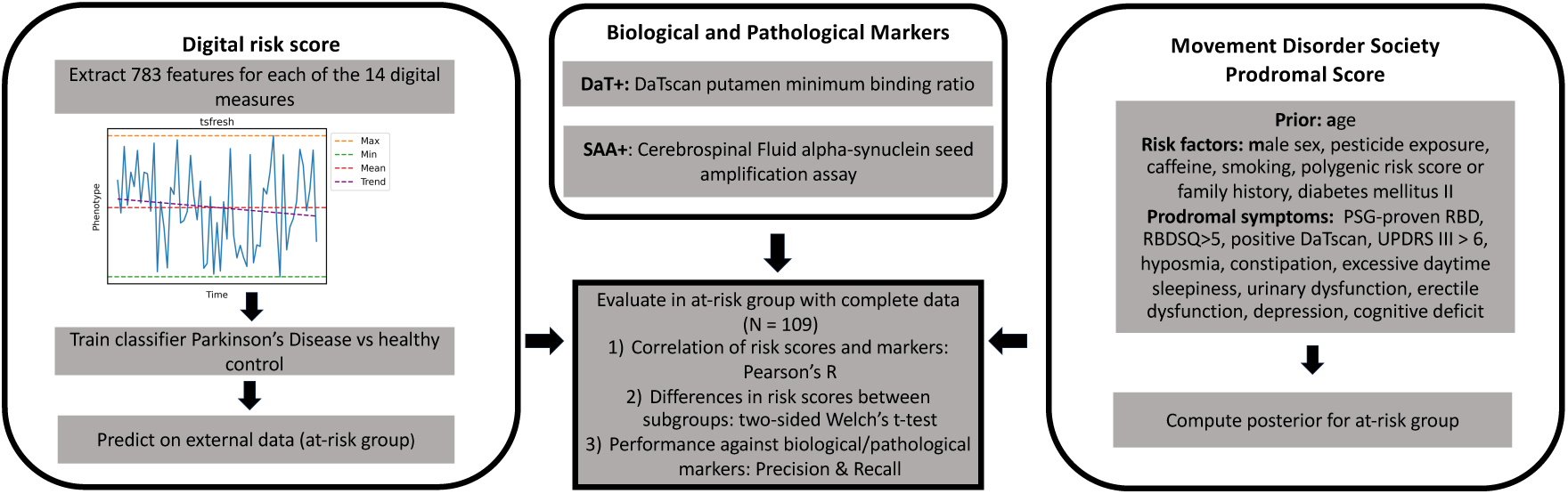
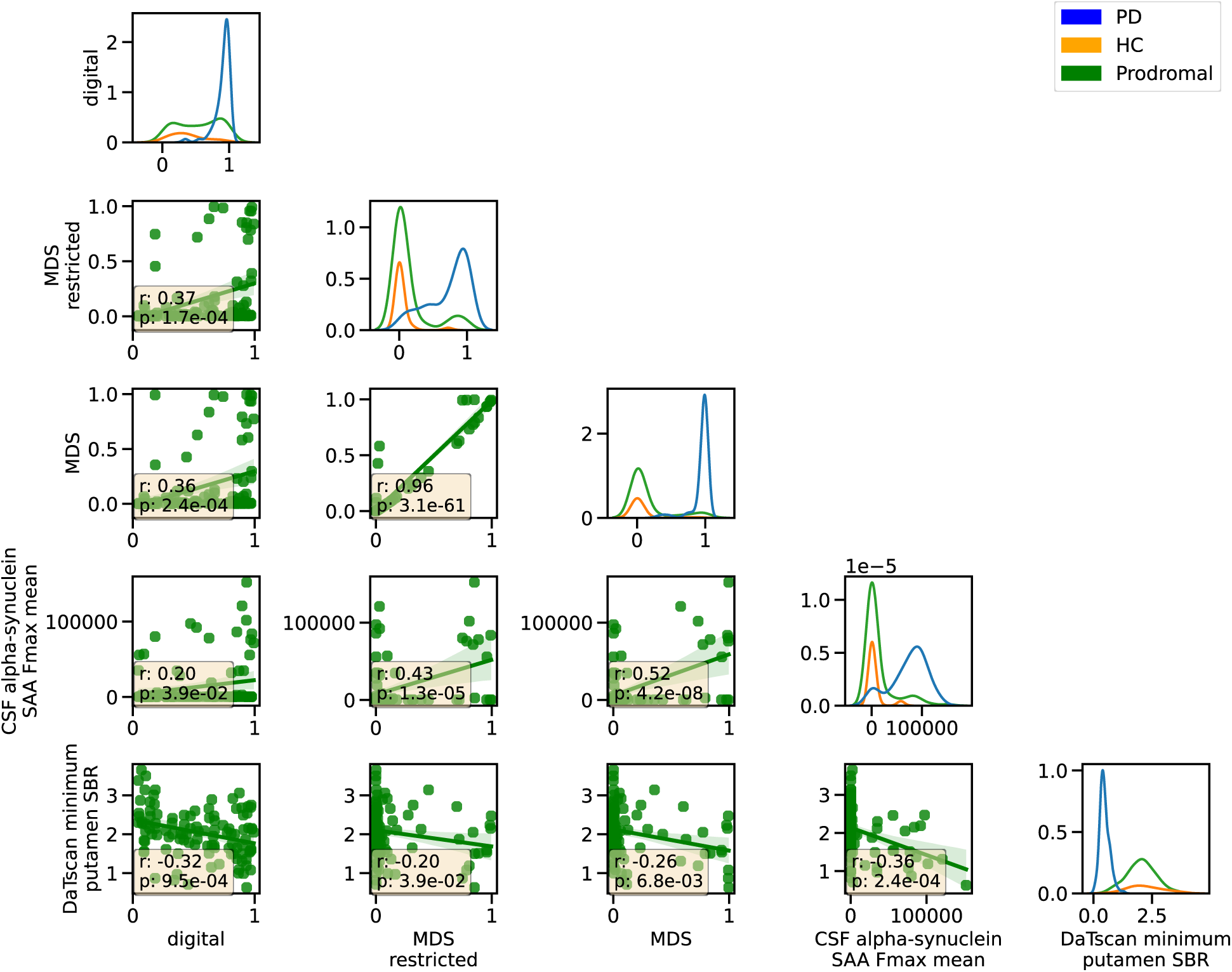
Digital risk correlates with prodromal score and biological markers A) Overview of analysis. Derivation of risk scores and biological and pathological markers. Illustration of performed tests and modelling. B) The relation between the different risk scores and biological markers is shown. On the diagonal, the distribution for each diagnostic group is displayed (PD: diagnosed Parkinson’s disease, HC: healthy control, Prodromal: at-risk cohort of genetic mutations carriers and individuals with prodromal symptoms). The scatterplot shows the relation between each pair of markers in the prodromal group (N = 109) with the text box displaying the Pearson r coefficient and the 0.05 FDR corrected p-value.

**Figure 3:**
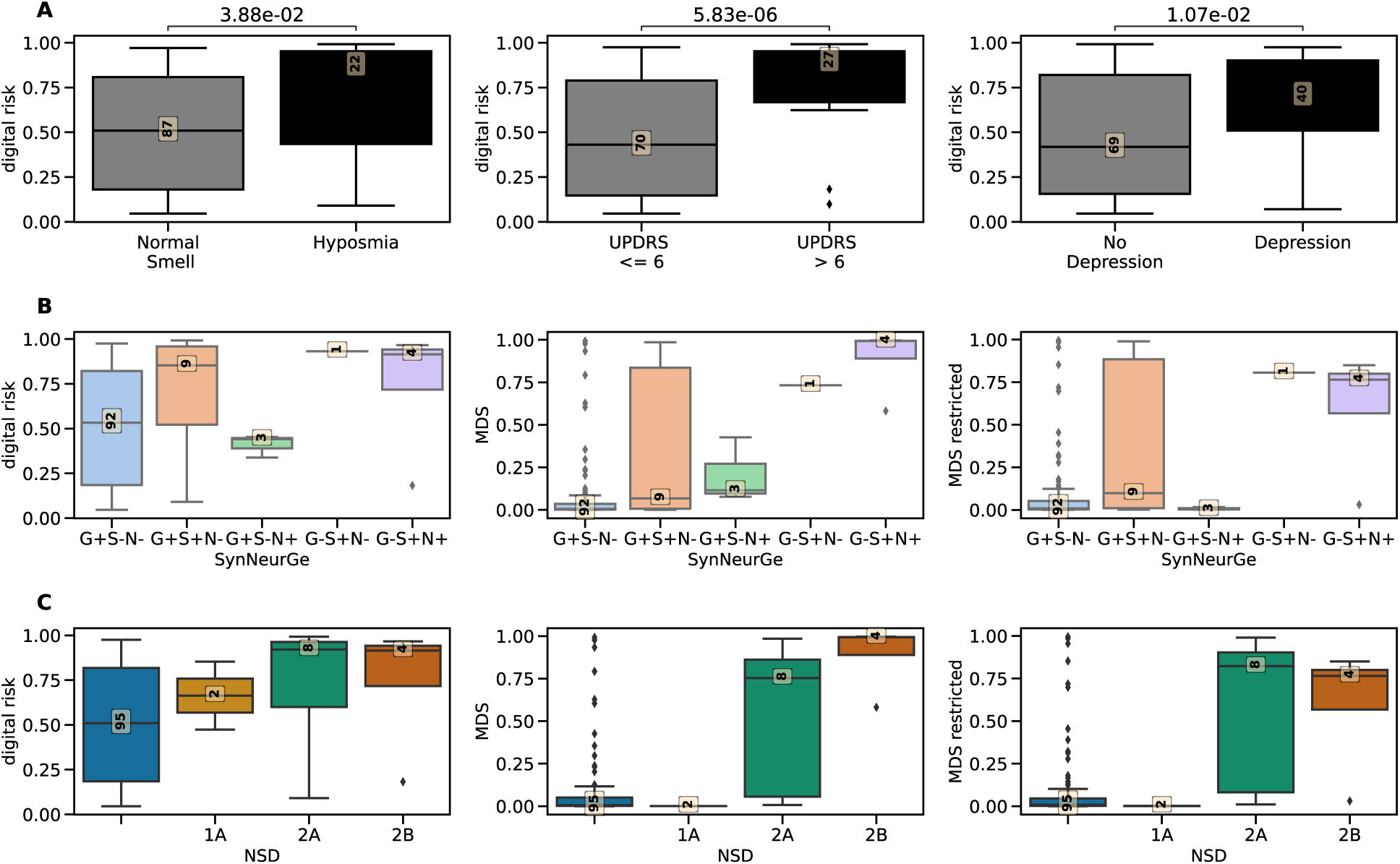
**Digital risk score is increased in individuals with known prodromal markers and biological classification groups** A) The boxplots show the difference in digital risk score between carriers and non-carriers (x-axis). The 0.05 FDR-corrected p-value from two-sided Welch t-test is shown. The yellow box presents the number of subjects in each group. This plot shows those prodromal markers and risk factors from the model included in Heinzel, Berg, Gasser, Chen, Yao, Postuma and Disease 2 that were significant after FDR-correction. A complete table with statistical results can be found in STable 6. B) The distribution of risk scores for the different biological groups defined by SynNeurGe(11) for the digital, the MDS, and the restricted MDS risk scores. C) The distribution of the risk score for the different biological stages defined by NSD(10) for the digital, MDS, and the restricted MDS risk scores.

Additionally, we computed a prodromal risk score using the model defined in the MDS research criteria (2) based on the longitudinal information on lifestyle, prodromal symptoms, and genetics (Figure 2A, STable 2). DaTscan being one of the most predictive but least performed investigations clinically, a restricted version was additionally computed without DaTscan information, better representing the real-world application. Our derived digital risk score was significantly correlated with the MDS risk score (r = 0.36, p-value = 2.43x10^-4^, N = 109, Figure 2B, STable 8) and the restricted model excluding DaTscan information (r = 0.37, p-value = 1.65x10^-4^, Figure 2B).

The digital risk score was further correlated with the minimum putamen SBR as derived from DaTscan (r = -0.32, p-value = 9.49x10^-4^) and the CSF alpha-synuclein SAA (r = 0.2, p-value = 3.9x10^-2^, Figure 2B). The MDS risk scores also correlated with these biological and pathological markers with the digital risk score showing stronger correlation with DaTscan and the MDS prodromal a stronger association with CSF alpha-synuclein SAA. Overall, the digital risk score can be assumed to capture information relevant to the biological and pathological risk for PD.

### Digital risk score is increased in individuals with hyposmia or subthreshold Parkinsonism

We investigated which known risk factors and prodromal symptoms implied significant differences in digital risk. The digital risk score captured significant differences for subthreshold parkinsonism (UPDRS III > 6) (p-value = 5.83x10^-6^), hyposmia (p-value = 3.88x10^-2^), and depression (p-value = 1.07x10^-2^) (Figure 3A, STable 9, STable 10).

Following the SynNeurGe biological definition of PD, our at-risk cohort had 92 genetically predisposed individuals (GP+S-N-), 9 genetic Parkinson’s type synucleinopathy (GP+S+N-), 3 Non-PD neurodegeneration (GP+S-N+), 4 sporadic PD (GP-S+N+), and 1 sporadic Parkinson’s type synucleinopathy (GP-S+N-) (Figure 3B). Our digital risk score was highest in individuals with sporadic PD (0.74±0.38) and lowest for groups without synucleinopathy (GP+S-N-: 0.52±0.32, GP+S-N+: 0.41±0.06) (Figure 3B). Similar results were observed for the MDS prodromal scores. Following the NSD staging system, 95 individuals are not assigned to any stage, 2 to 1A (S+N-C-), 8 to 2A (S+N-C+), and 4 to 2B (S+N+C+) (Figure 3C). This system does not allow for S- in presence of D+, which occurs in 4 individuals in our cohort, potentially due to the SAA being conducted at an earlier time point. The digital risk score increases with each stage whereas the MDS prodromal score shows a steep increase only from stage 2 (Figure 3C).

### Digital risk score as a sensitive screening tool

Due to a lack of future conversion information, we display the performance of each risk score against the combination of DaTscan positivity and SAA positivity. The number of true positives (TP), false positives (FP), true negatives (TN), and false negatives (FN) is shown alongside precision, recall, and F1 score.

Importantly, MDS includes information on DaTscan positivity in its model, hence, we included a restricted version without this information as well.

The digital risk score showed a bi-modal distribution for the at-risk group compared to a heavy-tailed one in the MDS model (Figure 2B). It identified 51.38% (N = 56) of the at-risk group as high risk whereas the MDS research criteria only flagged 8.26% (N = 9). Of the 109 subjects investigated with complete data available, 12.84% (N = 14) had positive SAA and 6.42% (N = 7) positive DaTscan (Figure 4A, Table 1). The digital risk model might thus misidentify several cases as high risk.

**Figure 4:**
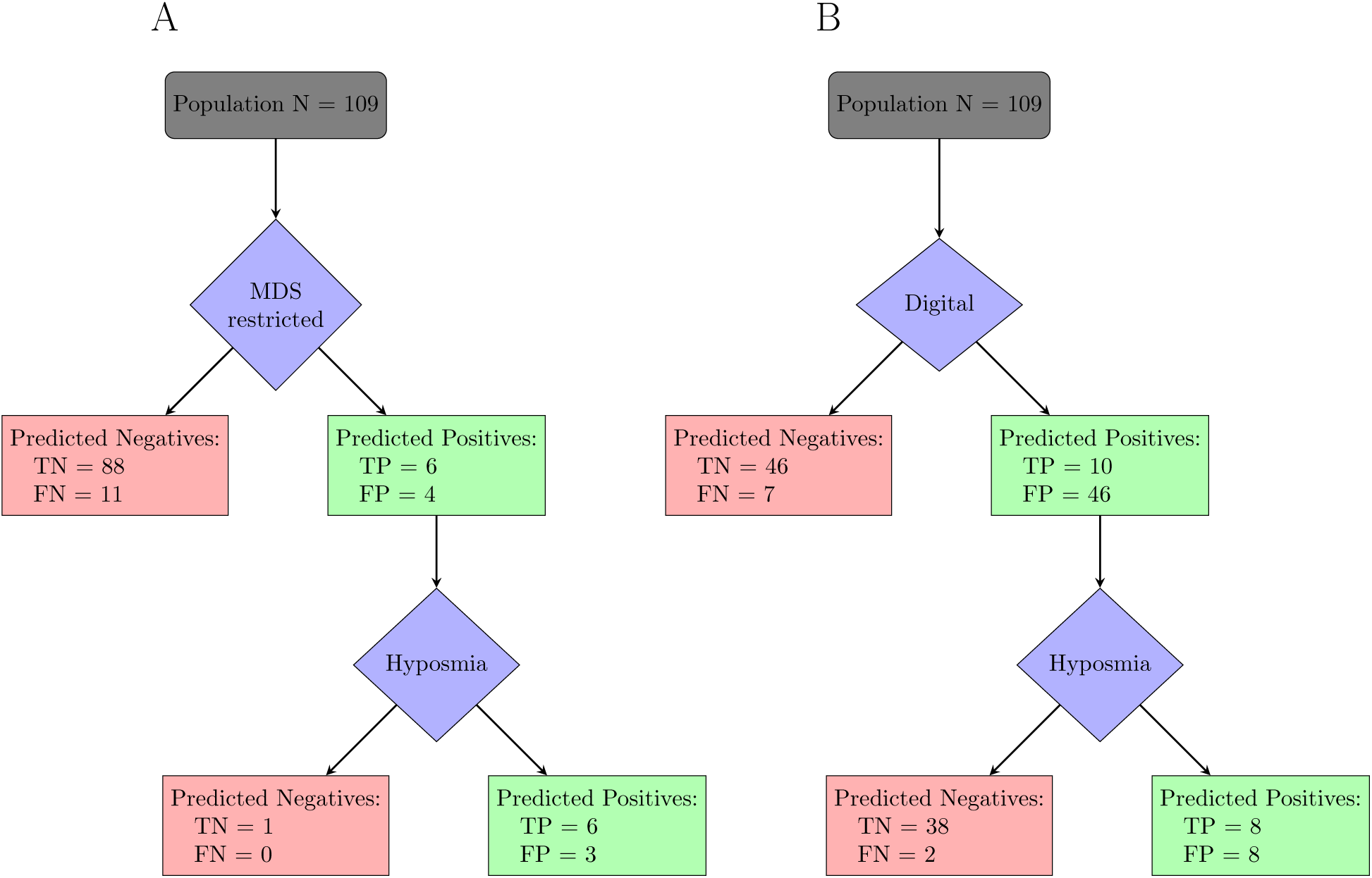
Digital risk score as a sensitive first screening tool in a sequence of testing The flowcharts displays a potential chaining of tests to identify people at risk of Parkinson’s disease. Here the tests are evaluated against presence of biological (DaTscan) or pathological (alpha-synuclein SAA) markers due to a lack of future phenoconversion information. A) Starting with the restricted MDS prodromal risk as defined by Heinzel, Berg, Gasser, Chen, Yao, Postuma and Disease 2, 10 people are sent for further examination of hyposmia. B) Starting with the most sensitive and accessible test, the digital risk score, 56 of the 109 individuals are sent for further testing to evaluate hyposmia which increases the specificity.

**Table 1:** The digital risk score sensitively identifies people with biological markers of Parkinson’s disease

|  | TN | FP | FN | TP | precision | recall | F1 score |
| --- | --- | --- | --- | --- | --- | --- | --- |
| MDS | 89 | 3 | 11 | 6 | 0.67 | 0.35 | 0.46 |
| MDS restricted | 88 | 4 | 11 | 6 | 0.6 | 0.35 | 0.44 |
| Hyposmia | 80 | 12 | 7 | 10 | 0.45 | 0.59 | 0.51 |
| digital | 46 | 46 | 7 | 10 | 0.18 | 0.59 | 0.27 |
| digital+hyposmia | 42 | 50 | 5 | 12 | 0.19 | 0.71 | 0.3 |
| SAA | 92 | 0 | 3 | 14 | 1 | 0.82 | 0.9 |
| DaTscan | 92 | 0 | 10 | 7 | 1 | 0.41 | 0.58 |

With no current gold-standard available for conversion, as no individual received a diagnosis of PD after digital data collection, we assessed the risk model performance against biological and pathological markers known to be altered in prodromal PD (8, 9, 20).

The digital model identified a higher number of the biological at-risk people than the MDS model (Table 1, STable 11). The digital risk score better identified SAA positivity (recall = 0.71) than DaTscan positivity (recall = 0.43). It showed recall equal or higher than the MDS prodromal models, however, had lower precision (Table 1, STable 11). Compared to hyposmia as determined by UPSIT test and medical records (STable 2), the digital model had equal recall for SAA positivity. Generally, hyposmia performed better or equal to the digital risk score. The two tests, however, identified distinct individuals for SAA positivity with a combined risk identifying 12 of the 14 SAA positive cases. This indicates a potential benefit in creating a combined risk score leveraging hyposmia and digital risk scores.

A potential application of the digital risk could be the first sensitive screening which is followed by more specific testing. All predicted positive cases from the digital risk model would be sent for further testing with hyposmia, with the final examination being performed with CSF alpha-synuclein SAA or DaTscan (Figure 4). In contrast, if the initial screening were performed with the restricted MDS prodromal risk score, 11 individuals with either DaT+ or SAA+ would be missed from the start.

The digital model could be biased towards identifying only those individuals already presenting with minor motor impairments (STable 12). The individuals that the digital model did not identify but that had either positive SAA or DaT, had lower maximum UPDRS III scores (mean = 1.86±2.27, N=7) than the ones correctly identified by the digital risk (mean = 15.7±11.33, N=10) (p-value = 3.74x10^-3^, dof = 10, Cohen’s d = 1.56). Hyposmia (p-value = 2.58x10^-3^) and the restricted MDS prodromal risk score (p-value = 2.15x10^-4^) showed this same bias towards individuals with higher UPDRS III scores being identified and those with lower being missed.

## DISCUSSION

We leveraged the 1.3-years of continuously collected smartwatch data from the PPMI cohort to derive a digital risk score. Individuals with known prodromal markers, subthreshold Parkinsonism or hyposmia, demonstrated an increased digital risk. The digital risk score showed a high sensitivity but at the same time concordance with MDS research criteria(2), CSF alpha-synuclein SAA, and DaTscan.

We have previously demonstrated the ability to identify those who will go on to receive a future diagnosis of PD using one week of accelerometer data (12). Here, we evaluated long-term digital markers derived from a multi-sensor device worn by individuals harboring genetic risk variants or prodromal markers for PD. We assessed the improvement in using the whole observation time versus the last week of data available and found a significant improvement for the long-term risk (p-value = 0.03, mean AUPRC difference = 0.1, SMethods). The addition of the PPG sensor allowed the extraction of sleep stages and vital signs, which significantly contributed to the digital risk (Figure 2&4, SMethods). The extended timeframe and measures thus contributed to an improved digital risk score.

With the digital risk correlating with neurological markers of PD, its relevance for early screening was further highlighted. Compared to the MDS research criteria without DaTscan information, the digital risk score had a higher recall for CSF alpha-synuclein SAA positivity (increase by 0.29) and DaTscan positivity (increase by 0.29). Previous research reported hyposmia as a good predictor for DaTscan positivity (21) with a recall of 0.96 and a precision of 0.14. In our dataset, hyposmia only achieved a recall of 0.57 and a precision of 0.18. This discrepancy could be attributed to a different method for ascertainment of hyposmia or population characteristics. Generally, hyposmia performed as well or better than the digital risk score. However, we noted that distinct individuals were identified for CSF alpha-synuclein SAA positivity by the two tests, with a combination identifying 85.71% of CSF alpha-synuclein SAA positive cases.

The digital risk score identified half of the at-risk group as high-risk. This high rate could relate to a high rate of false positives, placing a burden on healthcare systems to undertake additional screening tests, and anxiety for the individuals incorrectly identified as being at high-risk. Prior to additional invasive testing being undertaken based on digital risk, further clinical examination should be performed (including testing for hyposmia), aligning with current recruiting strategies for prodromal cohorts(22). The true rate of false positives remains to be determined due to the missing information of future phenoconversion in the current dataset. Although CSF alpha-synuclein SAA and DaTscan are good markers for the neuropathological changes associated with PD, they are not diagnostic tools (9). Future follow-up is thus important to assess the true predictive performance of the risk scores.

The accessibility of a digital risk score is much higher than for current prodromal models as it does not rely on the availability of DaTscan and other testing, which is generally only performed when PD is already suspected (23). DaTscan has previously been proposed to be used as a secondary test following a more sensitive and accessible one (24). Hyposmia is currently the most promising early screening marker for PD with the test being low-cost and easily accessible (21, 25). Here, we showed that a digital risk score could offer an alternative or addition to this test with passively collected digital data having the advantage of continuous and passive longer-term follow-up.

The primary limitations of this study relate to data availability and choice of methodology. Due to the Verily Study Watch only being introduced 10 years after the start of the PPMI study, for some individuals the different data modalities have been collected several years apart, limiting the comparability between modalities to indicate risk. With the digital risk being collected most recently, its risk detection performance could be attributed to individuals being potentially closer to phenoconversion. For example, the alpha-synuclein SAA is currently only available for baseline data. Regarding the risk calculations, the at-risk group currently only includes 28 individuals known to have subsequently received a diagnosis (converters) of which only seven had digital data available with six converting prior to data collection and one without a known conversion date, limiting the assessment of the true risk of developing PD. Our study was further limited to the derived features provided by Verily, the code for which is proprietary, limiting reproducibility in other cohorts. As sleep scores differ highly between devices and employed processing and have not reached the same performance as achieved with PSG (26), the included sleep features should be interpreted with this variability in mind. Finally, due to lack of an independent validation cohort, our findings remain to be replicated in the future.

## CONCLUSION

In conclusion, long-term digital monitoring can inform disease risk which relates to existing risk scores and the underlying biology of PD. A digital risk could serve as an initial screening test, followed by more specific tests for diagnostic confirmation, such as a DaTscan or CSF alpha-synuclein SAA measurement.

## Data Availability

This analysis used DaTscan and alpha-synuclein SAA results for at-risk participants, obtained from PPMI upon request after approval by the PPMI Data Access Committee. Data used in the preparation of this article were obtained in November 2022 from the Parkinson's Progression Markers Initiative (PPMI) database (www.ppmi-info.org/access-data-specimens/download-data), RRID:SCR_006431. Sequestered data was given access to October 2023.

## Acknowledgments

We thank all participants of the PPMI study, all investigators, and the Michael J. Fox Foundation.

## Author contributions

Conceptualization: AKS, CS Methodology: AKS Investigation: AKS Visualization: AKS Supervision: CS

Writing – original draft: AKS

Writing – review & editing: AKS, NH, KP, VEP, PB, CS

## Data and materials availability

This analysis used DaTscan and alpha-synuclein SAA results for at-risk participants, obtained from PPMI upon request after approval by the PPMI Data Access Committee. Data used in the preparation of this article were obtained in November 2022 from the Parkinson’s Progression Markers Initiative (PPMI) database (www.ppmi-info.org/access-data-specimens/download-data), RRID:SCR_006431. Sequestered data was given access to October 2023. For up-to-date information on the study, visit www.ppmi-info.org.

All associated code to reproduce the analyses performed here will be made publicly available upon publication (https://github.com/aschalkamp/DigitalPPMI).

## References

1. Fearnley JM, Lees AJ. Ageing and Parkinson’s disease: substantia nigra regional selectivity. Brain. 1991;114(5):2283–301.

2. Heinzel S, Berg D, Gasser T, Chen H, Yao C, Postuma RB, et al. Update of the MDS research criteria for prodromal Parkinson’s disease. Mov Disord. 2019;34(10):1464–70.

3. Bestwick JP, Auger SD, Simonet C, Rees RN, Rack D, Jitlal M, et al. Improving estimation of Parkinson’s disease risk-the enhanced PREDICT-PD algorithm. NPJ Parkinsons Dis. 2021;7(1):33.

4. Mahlknecht P, Gasperi A, Djamshidian A, Kiechl S, Stockner H, Willeit P, et al. Performance of the Movement Disorders Society criteria for prodromal Parkinson’s disease: A population-based 10-year study. Mov Disord. 2018;33(3):405–13.

5. Yilmaz R, Suenkel U, Team TS, Postuma RB, Heinzel S, Berg D. Comparing the Two Prodromal Parkinson’s Disease Research Criteria-Lessons for Future Studies. Mov Disord. 2021;36(7):1731–2.

6. Iranzo A, Lomena F, Stockner H, Valldeoriola F, Vilaseca I, Salamero M, et al. Decreased striatal dopamine transporter uptake and substantia nigra hyperechogenicity as risk markers of synucleinopathy in patients with idiopathic rapid-eye-movement sleep behaviour disorder: a prospective study [corrected]. Lancet Neurol. 2010;9(11):1070–7.

7. Eisensehr I, Linke R, Noachtar S, Schwarz J, Gildehaus FJ, Tatsch K. Reduced striatal dopamine transporters in idiopathic rapid eye movement sleep behaviour disorder. Comparison with Parkinson’s disease and controls. Brain. 2000;123 ( Pt 6):1155–60.

8. Chahine LM, Brumm MC, Caspell-Garcia C, Oertel W, Mollenhauer B, Amara A, et al. Dopamine transporter imaging predicts clinically-defined alpha-synucleinopathy in REM sleep behavior disorder. Ann Clin Transl Neurol. 2021;8(1):201–12.

9. Siderowf A, Concha-Marambio L, Lafontant DE, Farris CM, Ma Y, Urenia PA, et al. Assessment of heterogeneity among participants in the Parkinson’s Progression Markers Initiative cohort using alpha-synuclein seed amplification: a cross-sectional study. Lancet Neurol. 2023;22(5):407–17.

10. Simuni T, Chahine LM, Poston K, Brumm M, Buracchio T, Campbell M, et al. A biological definition of neuronal alpha-synuclein disease: towards an integrated staging system for research. Lancet Neurol. 2024;23(2):178–90.

11. Hoglinger GU, Adler CH, Berg D, Klein C, Outeiro TF, Poewe W, et al. A biological classification of Parkinson’s disease: the SynNeurGe research diagnostic criteria. Lancet Neurol. 2024;23(2):191–204.

12. Schalkamp A-K, Peall KJ, Harrison NA, Sandor C. Wearable movement-tracking data identify Parkinson’s disease years before clinical diagnosis. Nature Medicine. 2023.

13. Parkinson Progression Marker I. The Parkinson Progression Marker Initiative (PPMI). Prog Neurobiol. 2011;95(4):629–35.

14. Doty RL, Shaman P, Dann M. Development of the University of Pennsylvania Smell Identification Test: a standardized microencapsulated test of olfactory function. Physiol Behav. 1984;32(3):489–502.

15. Goetz CG, Tilley BC, Shaftman SR, Stebbins GT, Fahn S, Martinez-Martin P, et al. Movement Disorder Society-sponsored revision of the Unified Parkinson’s Disease Rating Scale (MDS-UPDRS): scale presentation and clinimetric testing results. Mov Disord. 2008;23(15):2129–70.

16. Stiasny-Kolster K, Mayer G, Schafer S, Moller JC, Heinzel-Gutenbrunner M, Oertel WH. The REM sleep behavior disorder screening questionnaire--a new diagnostic instrument. Mov Disord. 2007;22(16):2386–93.

17. Pedregosa F, Varoquaux G, Gramfort A, Michel V, Thirion B, Grisel O, et al. Scikit-learn: Machine Learning in Python. Journal of Machine Learning Research. 2011;12(85):2825—30.

18. Christ MB, N., Neuffer, J., Kempa-Liehr, A.W. Time Series {FeatuRe} Extraction on basis of Scalable Hypothesis tests (tsfresh {\textendash} A Python package). Neurocomputing. 2018;307:72–7.

19. Vallat R. Pingouin: statistics in Python. Journal of Open Source Software. 2018;3:1026.

20. Jennings D, Siderowf A, Stern M, Seibyl J, Eberly S, Oakes D, et al. Conversion to Parkinson Disease in the PARS Hyposmic and Dopamine Transporter-Deficit Prodromal Cohort. JAMA Neurol. 2017;74(8):933–40.

21. Jennings D, Siderowf A, Stern M, Seibyl J, Eberly S, Oakes D, et al. Imaging prodromal Parkinson disease: the Parkinson Associated Risk Syndrome Study. Neurology. 2014;83(19):1739–46.

22. Mahlknecht P, Marini K, Werkmann M, Poewe W, Seppi K. Prodromal Parkinson’s disease: hype or hope for disease-modification trials? Transl Neurodegener. 2022;11(1):11.

23. de la Fuente-Fernandez R. Role of DaTSCAN and clinical diagnosis in Parkinson disease. Neurology. 2012;78(10):696–701.

24. Postuma RB. Dopaminergic Imaging and Prodromal Parkinson Disease: A Key Biomarker Arrives. JAMA Neurol. 2017;74(8):901–3.

25. Ross GW, Petrovitch H, Abbott RD, Tanner CM, Popper J, Masaki K, et al. Association of olfactory dysfunction with risk for future Parkinson’s disease. Ann Neurol. 2008;63(2):167–73.

26. Miller DJ, Sargent C, Roach GD. A Validation of Six Wearable Devices for Estimating Sleep, Heart Rate and Heart Rate Variability in Healthy Adults. Sensors (Basel). 2022;22(16).

