## Supplemental Material for "Digital risk score sensitively identifies presence of α-synuclein aggregation or dopaminergic deficit"

### Supplemental Methods

#### Exploration of digital risk score

We explored the effect of different feature sets on the digital risk score as well as searched for the best performing Machine Learning model.

##### 1.1 Machine Learning Model

We explored the performance of various ML models to identify PD from healthy controls using the digital timeseries features. We compared logistic regression with elastic net penalty to random forests, support vector machines with polynomial kernel, and support vector machines with radial basis functions (eTable 7). This served to identify whether non-linear associations are important for model performance and ensure good performance overall. All models were trained as outlined in the main manuscript in nested 5-fold cross validation where the inner loop performed gridsearch to identify the best hyperparameters. Performance was compared as area under precision recall curve (AUPRC) across the five outer folds. Receiver operator curves and precision recall curves are presented as well (eFigure 2). Compared to logistic regression, none of the models significantly outperformed it after 0.05 Bonferroni-correction (p-value = 0.99, p-value = 0.53, p-value = 0.02). We chose logistic regression over the other models as it showed similar performance while being the simplest and most interpretable one.

##### 1.2 Feature sets

We explored how a restriction to specific feature sets affects performance. For this, we trained three models. One restricted to physical activity features, one to vital signs, and one to sleep. We compared their performance to that of the combined model (eFigure 3). The combined model outperformed all, and significantly did so for physical activity (AUPRC: 0.94±0.004, p-value = $2.09 \times 10^{-3}$) and vital signs (AUPRC: 0.79±0.03, p-value = $1 \times 10^{-6}$). The model restricted to sleep features (AUPRC: 0.95±0.02, p-value = 0.09) performed best and on-par with the combined model (AUPRC: 0.96±0.01). The one based on vital signs did not perform better than the baseline model trained on age and sex alone (AUPRC: 0.8±0.04).

##### 1.3 Considered time-frame

We analysed how the digital risk score would perform if restricted to one week of data as compared to the model using the whole observation time of 1.3 years on average. For this, we identified the last hour when data was recorded for each subject (based on step count information) and extracted the data up to seven days before. We then applied tsfresh as before, obtaining 783 features per timeseries and fitted the model identifying PD from healthy controls just as detailed in the main manuscript. The resulting model performed worse than the one trained on the whole observation period (t = 2.51, dof = 8, p-value = 0.04, 95% CI = [0.01,0.2]) (eFigure 4).

### Supplemental Tables

STable 1: Derived digital markers as provided by Verily

| modality | category | sensors | #features | features | model |
|---|---|---|---|---|---|
| physical activity | ambulatory | 3-axis accelerometer | 1 | hourly walking minutes | 2-class classifier (walk/run vs other) trained on 215000 hours of self-report labelled free-living data from 1800 adult subjects with out-of sample performance of 87% |
| | step | 3-axis accelerometer | 1 | hourly step count | frequency-based model validated against ankle-worn gait monitor on 329 days of free-living data of 75 adult subjects with 18% mean absolute error |
| sleep | sleep onset/offset | accelerometer, PPG | 4 | sleep efficiency, number of awakenings, total sleep time, wake after sleep onset | algorithm trained on PPG and ECG validated against majority vote of three wearables on 176 nights in home setting of 50 adult subjects with median absolute error of sleep onset of 6 minutes and 9 minutes for sleep offset |
| | sleep stages | accelerometer, PPG | 4 | REM, NREM, light NREM, deep NREM | algorithm trained on PPG and ECG validated against majority vote of three wearables on 176 nights in home setting of 50 adult subjects with an overall accuracy of 70% [1] |
| vital signs | pulse rate | PPG | 1 | total mean pulse rate per hour | algorithm from ADI validated against heart rate of ECG on one to two hours of in-clinic data of 50 adult subjects with a mean absolute error of 10.7 beats per minute ADI2023 |
| | pulse rate variability | PPG | 3 | mean, median and variance of hourly RMSSD | algorithm measuring RMSSD of interbeat intervals validated against 510000 wearables for ECG on 200 days of free-living data of 50 adult subjects with mean absolute error of |

9.9ms [2]

The different hourly statistics as derived from the smartwatch data are described. This information is taken from the accompanying documents on PPMI LONI. PPG: Photoplethysmography, ECG: Electrocardiography, RMSSD: root mean square of successive differences between normal heartbeats, REM: rapid eye movement

STable 2: Risk factors and Prodromal Markers

| | | |
|---|---|---|
| risk factors | | |
| | age | age at data retrieval date: 01.10.2021 |
| | sex | male |
| | pesticide exposure | FOUND questionnaire whether occupational exposure |
| | non-use of caffeine | FOUND questionnaire less than 6 cups of tea or 3 cups of coffee weekly |
| | never smoke | FOUND questionnaire not ever smoked regularly |
| | previous smoke | FOUND questionnaire ever smoked regularly and not smoke currently |
| | current smoke | FOUND current regular smoker |
| | Physical inactivity | |
| | 1st degree relative with PD | mother, father, or sibling with PD diagnosis (only used when PRS unavailable) |
| | PRS | Polygenic risk score calculated with Nalls, Pankratz [3],<br>low if in lowest quartile, high if in highest quartile |
| | diabetes mellitus type II | medical condition log searched for '(?!.*pre)(?!.*borderline)((.*(II\|2\|two).*Diabet.*)\|.*Diabet.*type.*(II\|2\|two).*)' |
| prodromal markers | | |
| | proven RBD | medical condition log searched for '.*(REM behavi\|RBD\|Rapid Eye).*' or listed under confirmed RBD in analytic dataset |
| | RBD test | ever scored higher than 5 on RBDSQ |
| | positive DaTscan | visual inspection of DaTscan abnormal or minimum putamen SBR 2std away from healthy control mean |

<table>
<tr><td></td><td>subthreshold parkinsonism</td><td>ever UPDRS III score excluding postural and kinetic tremor above 6</td></tr>
<tr><td></td><td>olfactory loss</td><td>medical condition log searched for '.*(hyposmia|anosmia).*' or listed under confirmed hyposmia in analytic dataset or ever scored below 1.5 std from age and sex matched mean [4]</td></tr>
<tr><td></td><td>constipation</td><td>medical condition log searched for '.*constipation.*' OR UPDRS I 1.11 > 1</td></tr>
<tr><td></td><td>excessive daytime sleepiness</td><td>medical condition log searched for '.*sleepiness.*' OR UPDRS I 1.13 > 1</td></tr>
<tr><td></td><td>urinary dysfunction</td><td>medical condition log searched for '(?!fecal).*incontinence.*' OR UPDRS I 1.10 > 1</td></tr>
<tr><td></td><td>orthostatic hypotension</td><td>medical condition log searched for '.*hypotension.*' OR UPDRS I 1.12 > 1</td></tr>
<tr><td></td><td>erectile dysfunction</td><td>medical condition log searched for '.*erectile.*' OR SCOPA autonome 22 > 1</td></tr>
<tr><td></td><td>depression</td><td>medical condition log searched for '.*(anxiety|depression).*' OR UPDRS I 1.3 > 1</td></tr>
<tr><td></td><td>cognitive deficit</td><td>ever cognitive categorisation listed as mild impairment or dementia</td></tr>
</table>

We describe the process of obtaining risk and prodromal markers from PPMI data. The selection of markers was taken from Heinzel, Berg [5].

#### STable 3: Study Cohort

Demographic and prodromal marker information for the PD, healthy control, and the different at-risk groups.

#### STable 4: Extracted digital timeseries features

The 783 feature extracted with tsfresh for each of the 14 digital measures are shown.

#### STable 5: Hyperparameters for Machine Learning models in gridsearch

| | Logistic regression | Polynomial Support Vector Machine | RBF Support Vector Machine | Random Forest |
|---|---|---|---|---|
| Penalty | Elastic net | | | |

| | | | | |
|---|---|---|---|---|
| C | np.logspace(1, 4, 5) | np.logspace(1, 4, 5) | np.logspace(1, 4, 5) | |
| L1-L2 ratio | np.linspace(0, 1, 5) | | | |
| Number of estimators | | | | [50,125,200] |
| Maximum depth | | | | [15,57,100] |
| degree | | [3,4,5] | | |

#### STable 6: Evaluation Cohort

**The at-risk group on which the digital risk score is evaluated is presented with proportion of prodromal markers present, mean age, and sex information.**

#### STable 7: Significant differences in digital markers between groups

The mean value per residual mean digital marker corrected for age and sex is shown for the healthy controls, the diagnosed PD, and the prodromal GBA, LRRK2, SNCA, hyposmia, RBD, and DaTscan positive cases together with the sample size per group. We show the statistics of the two sided T-test as the t-statistic and p-value. The digital markers are here the mean over the whole observation time for each individual.

#### STable 8: Correlation of risk scores and biological measures

We investigated the correlation of the risk scores and the biological measures with Pearson correlation. The statistics for each pair are shown with the sample size (n), r coefficient, 95% Confidence Interval (CI), uncorrected p-value, Bayes Factor (BF) 10, and power. After 0.05-Bonferroni adjustment, a p-value < 0.005 is significant.

#### STable 9: Differences in risk scores between risk factor and prodromal symptom carriers

The results of the two-sided T-tests are shown as the t-statistic, p-value, and number of individuals for each pair of risk factor/prodromal symptom and risk score.

#### STable 10: Mean risk score for each at-risk group

**For each at-risk group (LRRK2, GBA, hyposmia, RBD, DaT+, SAA+) we show the mean and standard deviation of the MDS, restricted MDS, and digital risk score.**

#### STable 11: Performance of the risk scores/markers in identifying biological/pathological risk

**The table displays the performance analysis of different risk scores or presence of prodromal markers for identifying SAA positivity, DaTscan positivity, or their combination. This is measured in true negatives (TN), false negatives (FN), true positives (TP), false positives (FP), as well as precision, recall, and F1 score.**

#### STable 12: Differences in UPDRS III between individuals at biological and pathological risk identified and missed by digital risk score

We report the statistical results of the comparison between false negatives and true positives for UPDRS III scores with two-sided Welch t-tests. We report the t-statistic, degrees of freedom, p-value, 95% Confidence Interval, and cohen’s d. This is reported for the comparison between the digital risk, restricted prodromal risk, and hyposmia with SAA or DaTscan positivity as the true outcome.

### Supplemental Figures

#### SFigure 1: Mean digital markers are affected in people diagnosed with PD

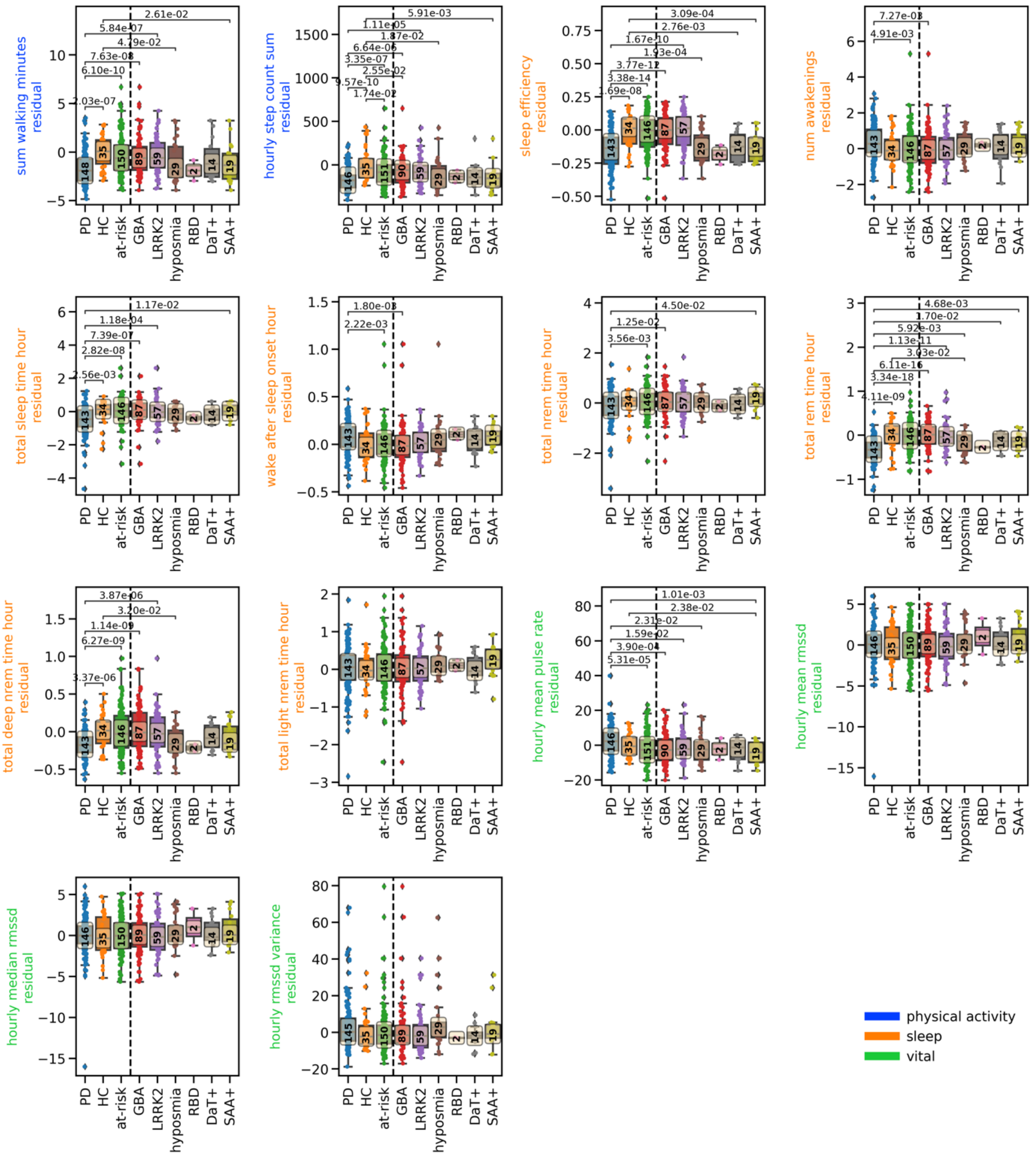

The boxplots show the residual overall mean adjusted for age and sex with parameters learned from a linear regression on the healthy controls. The overall mean is computed over the whole observation time per subject for each group for each digital marker. The boxplots depict the group median and quartiles per group with the whiskers showing the Q3+1.5 interquartile range (IQR) and Q1-1.5 IQR (Parkinson's disease cases: PD; healthy controls: HC; carriers of genetic risk alleles or prodromal symptoms without a diagnosis of PD: *GBA, SNCA, LRRK2*, olfactory loss, PSG-proven RBD, positive DaTscan, positive SAA, union of these: at-risk). The number in the yellow box indicates the number of individuals per group. Group differences were calculated with two-sided T-test between PD and HC to each of the prodromal groups. Asterisks show significant differences after 0.05 Bonferroni-correction.

#### SFigure 2: Significant predictors of digital risk model

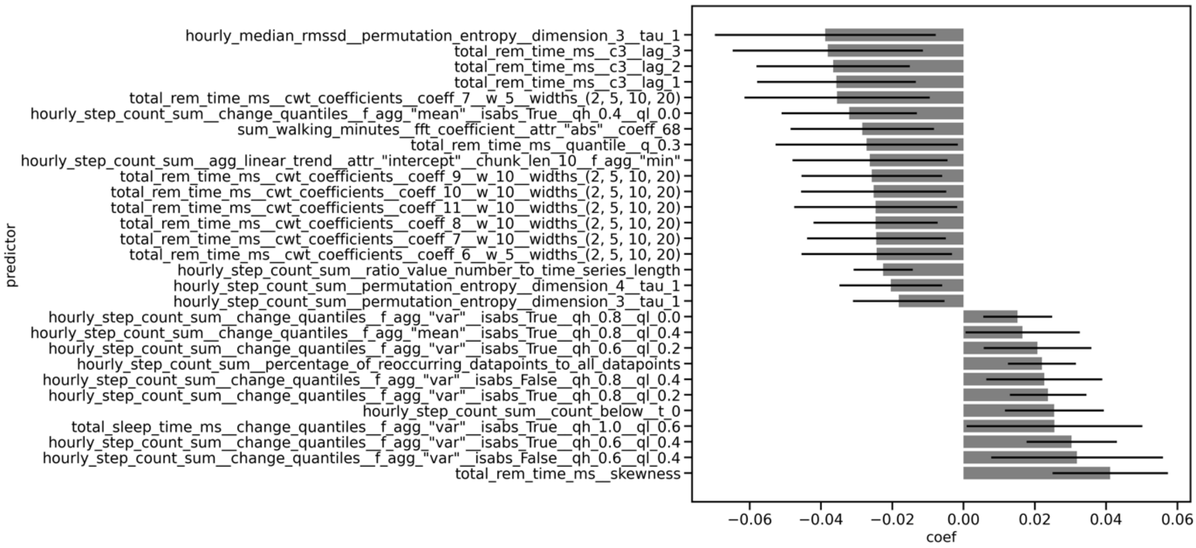

The predictors consistently and significantly selected across folds are shown with their mean effect size across folds and the 95% Bonferroni-corrected Confidence Interval. Significance across folds was determined with a one-sample ttest of the coefficients across folds with 0.05 Bonferroni-corrected significance level.

#### SFigure 3: Performance of digital risk models with different machine learning models

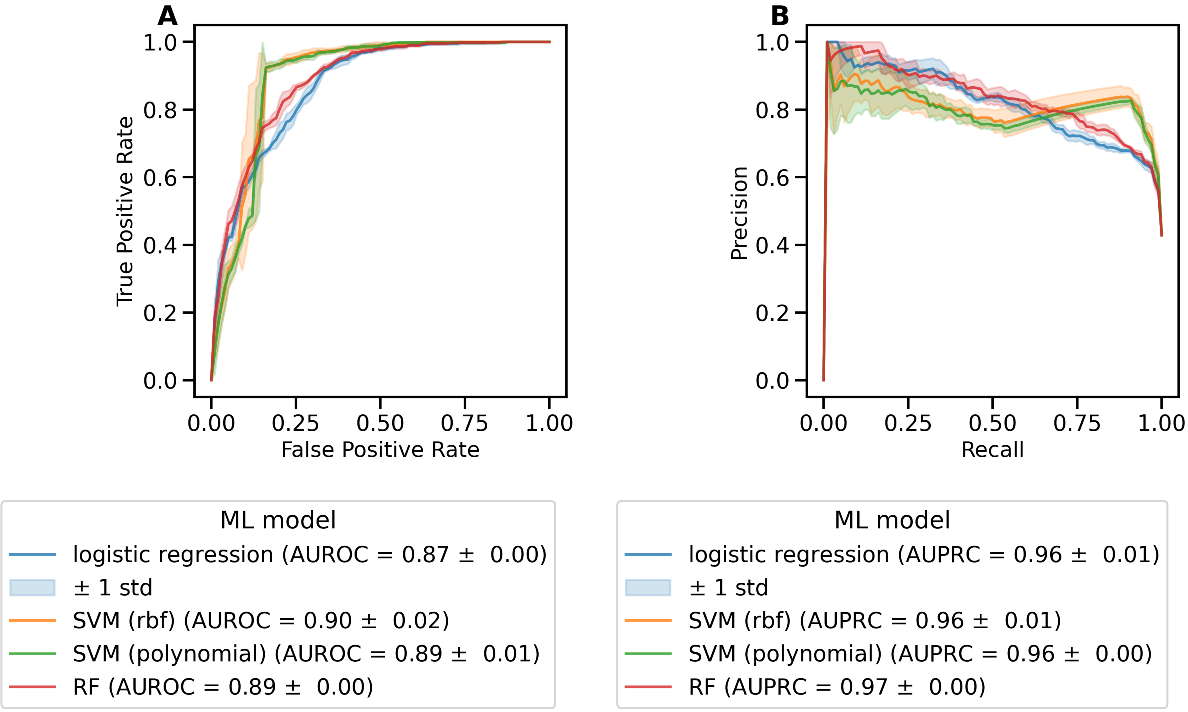

The performances for the digital risk score models is shown using different machine learning models. The A) receiver operator curve and the B) precision-recall curve are shown as the mean on the outer 5-folds of the nested cross-validation. The shaded area displays the standard deviation. For each classifier, the legend shows the mean A) area under receiver operator curve,  B) area under precision-recall curve with the standard error.

#### SFigure 4: Performance of digital risk models with different feature subsets

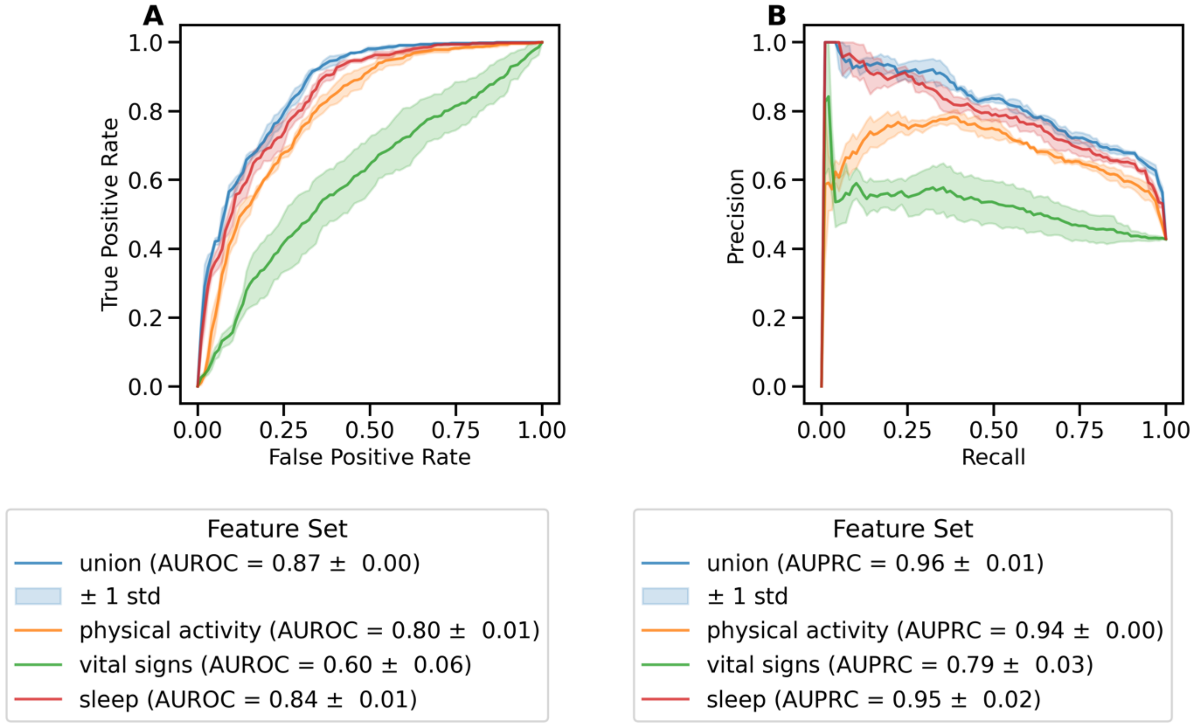

The performances for the digital risk score models is shown using different feature subsets where the training data was restricted to the digital features assigned to physical activity, sleep, or vital signs. The union of all is also displayed.. The A) receiver operator curve and the B) precision-recall curve are shown as the mean on the outer 5-folds of the nested cross-validation. The shaded area displays the standard deviation. For each classifier, the legend shows the mean A) area under receiver operator curve,  B) area under precision-recall curve with the standard error.

#### SFigure 5: Performance of digital risk models with different time-frames

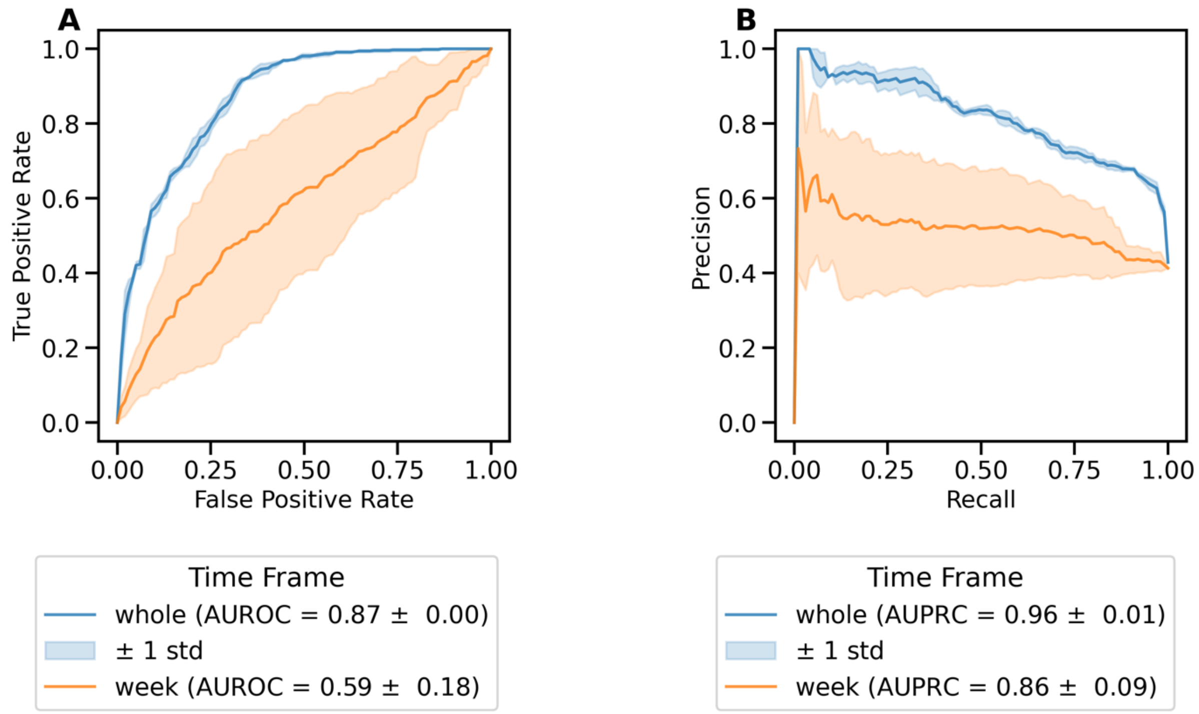

The performances for the digital risk score models is shown using different time-frames, either the whole observation period or the last week of available data. The A) receiver operator curve and the B) precision-recall curve are shown as the mean on the outer 5-folds of the nested cross-validation. The shaded area displays the standard deviation. For each classifier, the legend shows the mean A) area under receiver operator curve, B) area under precision-recall curve with the standard error.

### References

1. Sridhar, N., et al., *Deep learning for automated sleep staging using instantaneous heart rate.* NPJ Digit Med, 2020. **3**: p. 106.
2. Billman, G.E., *Heart rate variability - a historical perspective.* Front Physiol, 2011. **2**: p. 86.
3. Nalls, M.A., et al., *Large-scale meta-analysis of genome-wide association data identifies six new risk loci for Parkinson's disease.* Nat Genet, 2014. **46**(9): p. 989-93.
4. Brumm, M.C., et al., *Updated Percentiles for the University of Pennsylvania Smell Identification Test in Adults 50 Years of Age and Older.* Neurology, 2023. **100**(16): p. e1691-e1701.
5. Heinzel, S., et al., *Update of the MDS research criteria for prodromal Parkinson's disease.* Mov Disord, 2019. **34**(10): p. 1464-1470.
